# Development and Evaluation of Artificial Intelligence–Assisted Decision Support System for Public Health Emergency Classification and Escalation in Kenya

**DOI:** 10.64898/2026.07.07.26357475

**Authors:** Mark Nanyingi, Eric Osoro, Geoffrey H. Siwo, Isaac Ngere, Samuel Kadivane, James Magige, Joseph Kamau, Shreya Jain, Bryan O. Nyawanda, Joseph Njoroge, Ian Njeru, Kadondi Kasera, Victoria Kanana, Kamene Kimenye

## Abstract

**Background:** Timely assessment, classification, and escalation of public health events are essential for effective outbreak response, yet decision-making after event detection remains challenging because of fragmented guidance and variable interpretation of escalation criteria.To strengthen public health emergency management, Kenya developed the Decision-Making Tool for Public Health Emergencies (DMT-PHE), a framework for event assessment, classification, notification, and escalation. An artificial intelligence (AI)-enabled version, the DMT-PHE AI Agent, was subsequently developed to operationalize the framework through decision support. This study describes the development of the DMT-PHE AI Agent and evaluates its performance, usability, safety, and user acceptability.

**Methods:** The DMT-PHE AI Agent was developed using a retrieval-augmented generation architecture supported by a curated knowledge base derived from the validated DMT-PHE framework and related public health guidance. A simulation-based pilot evaluation was conducted among 11 public health professionals who independently assessed three standardized outbreak scenarios. AI-generated recommendations were compared with expert-defined gold standards. Outcomes included concordance, response-action coverage, citation performance, safety, usability, and user acceptability.

**Results:** Thirty-three scenario evaluations were completed. The AI Agent achieved an overall weighted concordance score of 0.924, with exact agreement of 90.9% for Public Health Events of Initially Unknown Etiology, 81.8% for Rift Valley fever, and 90.9% for Mpox. Citation support was provided in 78.8% of interactions, with no incorrect citations or major safety concerns identified. The mean System Usability Scale score was 85.2, while participants reported high trust (4.27/5), contextual relevance (4.55/5), and perceived time savings (4.82/5).

**Conclusions:** The DMT-PHE AI Agent demonstrated that a nationally validated public health emergency decision framework can be successfully translated into an AI-enabled decision-support system. These findings provide early evidence that AI can augment public health emergency decision-making by delivering structured, transparent, and context-specific recommendations while maintaining human oversight, offering a practical model for operationalizing national public health guidance.

## Introduction

Public health emergencies continue to pose substantial threats to global health security despite major investments in disease surveillance, laboratory networks, workforce development, and emergency preparedness systems. While advances in indicator-based surveillance, event-based surveillance, genomic surveillance, and epidemic intelligence have strengthened the early detection of public health threats, delays frequently occur after signals are identified, during event assessment, risk characterization, classification, escalation, and initiation of response activities. These post-detection decision-making processes are critical determinants of outbreak control because delays in recognizing the significance of an event or initiating appropriate public health action can result in continued transmission, increased morbidity and mortality, and wider socioeconomic disruption (1–7)

The International Health Regulations (IHR 2005) provide the principal global framework for assessing, notifying, and managing events that may constitute a Public Health Emergency of International Concern (1). More recently, the 7-1-7 framework has emphasized the importance of timely detection, notification, and response by proposing measurable targets for outbreak performance, namely detection within seven days of emergence, notification and initiation of investigation within one day of detection, and implementation of effective response measures within seven days (4,5). Although these frameworks provide clear expectations for public health action, they rely heavily on the ability of frontline practitioners to interpret multiple guidance documents, synthesize evolving epidemiological information, assess event significance, and determine appropriate escalation pathways. Consequently, the transition from surveillance information to operational decision-making remains a persistent challenge in many public health systems (2,3).

Artificial intelligence (AI) has emerged as a promising technology for strengthening health systems through enhanced data analysis, prediction, and decision support. Within public health, most AI applications have focused on disease surveillance, epidemic intelligence, outbreak forecasting, risk assessment, and signal detection, enabling earlier identification of emerging health threats (6–8). Comparatively less attention has been directed toward AI-assisted support for operational public health emergency management functions, including event classification, escalation, notification, coordination, and response planning. Yet these activities represent critical decision points that determine whether surveillance information is translated into timely and appropriate public health action (2,6,9,10)

Kenya provides an important setting in which to examine this implementation gap. The country has made substantial investments in public health surveillance, digital health information systems, laboratory networks, field epidemiology training, emergency operations capacity, and One Health coordination. These investments have strengthened the detection and reporting of priority diseases and public health threats at national and subnational levels(11–13). However, as in many health systems, the availability of surveillance data does not automatically ensure timely, consistent, and standardized decisions on event classification, notification, escalation, and response activation (1–5).Public health practitioners may still need to interpret multiple technical guidelines, escalation thresholds, disease-specific protocols, and coordination pathways under conditions of uncertainty and time pressure. This creates a critical post-detection gap between the generation of surveillance signals and the initiation of coordinated public health action.

Kenya and the wider East African region provide a compelling context for such decision support because they experience recurrent outbreaks of epidemic-prone, emerging, and re-emerging pathogens that require rapid assessment, escalation, and multisectoral coordination. In Kenya, recent and historical outbreaks have included cholera, dengue, chikungunya, Rift Valley fever, yellow fever, mpox, measles, and other priority public health threats, with outbreak reports documented across multiple counties between 2007 and 2022(14,15) The 2022 yellow fever outbreak in Isiolo County and the 2018, 2021 Rift Valley fever outbreak in northern Kenya illustrate the need for timely recognition, risk classification, laboratory confirmation, One Health coordination, and escalation of public health action(16,17).Kenya’s neighbours have also experienced major viral haemorrhagic fever events, including Sudan virus disease in Uganda and Marburg virus disease outbreaks in Tanzania and Rwanda, underscoring the regional risk posed by cross-border mobility, zoonotic spillover, delayed detection, and complex response coordination(18–20).These experiences highlight that surveillance and laboratory confirmation alone are insufficient unless accompanied by clear, standardized, and accessible decision support for determining when and how events should be classified, notified, escalated, and managed.

Recognizing this post-detection decision-making gap, the Kenya National Public Health Institute (KNPHI) developed the Decision-Making Tool for Public Health Emergencies (DMT-PHE), a standardized framework to guide assessment, classification, escalation, notification, and management of public health events across the Kenyan health system(21). The framework integrates the International Health Regulations (2005), national public health emergency management policies, Public Health Emergency Operations Centre (PHEOC) procedures, and One Health principles into a structured decision pathway for managing infectious disease outbreaks, zoonotic events, public health events of initially unknown etiology, chemical incidents, radiological emergencies, and other public health threats (1)

To improve accessibility and operational application of the framework, an AI-enabled decision-support system, the DMT-PHE AI Agent, was subsequently developed. The system was designed to operationalize the DMT-PHE framework through an interactive decision-support interface capable of assisting users with event assessment, classification, escalation, notification, and recommended response actions. By embedding nationally validated decision logic within an AI-assisted platform, the DMT-PHE AI Agent seeks to transform a static guidance framework into a source of real-time decision intelligence while maintaining human oversight and alignment with nationally approved public health emergency management procedures(9,10,22–25)

Although recent advances in generative AI have accelerated interest in AI-assisted decision support across healthcare, evidence regarding its application within operational public health emergency management remains limited, particularly in low- and middle-income countries. Furthermore, international guidance emphasizes that AI systems intended to support health decision-making should undergo rigorous early-stage evaluation to assess technical performance, usability, safety, transparency, and operational relevance before deployment in routine practice(22,23,25)

We describe the development of the DMT-PHE AI Agent and present the pilot evaluation of an AI-enabled decision-support system designed to operationalize a nationally validated public health emergency decision framework among public health professionals in Kenya

## Methods

### Study design

This study formed part of a broader multi-stage initiative to develop, validate, operationalize, and evaluate a standardized decision-support framework for public health emergencies in Kenya. The initiative began with the development of the Decision-Making Tool for Public Health Emergencies (DMT-PHE). The DMT-PHE was developed through an iterative consultative process involving national and county public health stakeholders and was subsequently refined through technical review and validation exercises to ensure alignment with the International Health Regulations (2005), Kenya’s public health emergency management architecture, Public Health Emergency Operations Centre (PHEOC) procedures, and One Health principles (1,21,26–28)

Following validation of the DMT-PHE framework, an artificial intelligence-assisted decision-support system, the DMT-PHE AI Agent, was developed to operationalize the framework through an interactive conversational interface. The AI Agent was designed to transform a static decision-support framework into a real-time decision-support system capable of assisting public health practitioners with event assessment, classification, escalation, notification, and identification of recommended response actions.

### Development of the Decision-Making Tool

The Decision-Making Tool for Public Health Emergencies (DMT-PHE) was conceptualized by the Kenya National Public Health Institute (KNPHI) through the Tackling Deadly Diseases in Africa Programme II (TDDAP2) as a standardized national framework to support consistent assessment, classification, escalation, notification, and response to public health events. The initiative sought to harmonize decision-making across national and subnational levels while strengthening implementation of the International Health Regulations (2005) and Kenya’s public health emergency management architecture (1,21,26)

Development followed a structured, iterative, and consultative process involving multidisciplinary experts from KNPHI, the Ministry of Health, county governments, epidemiology and disease surveillance programmes, Public Health Emergency Operations Centres (PHEOCs), laboratory services, veterinary and environmental health sectors, academia, and implementing partners. Existing national policies, emergency preparedness and response plans, disease-specific technical guidelines, and international guidance were systematically reviewed to identify the critical decision points required for timely assessment, risk classification, escalation, notification, coordination, and response to public health events (1,13,27–29)

Successive drafts of the DMT-PHE were refined through expert technical review and stakeholder consultation to ensure consistency with national emergency management structures, operational workflows, and One Health coordination mechanisms. The resulting framework provides standardized decision pathways for a broad spectrum of hazards, including infectious disease outbreaks, zoonotic diseases, public health events of initially unknown etiology, chemical incidents, radiological emergencies, environmental hazards, and other events requiring coordinated multisectoral response(1)

The finalized toolkit underwent national validation through a multi-stakeholder technical workshop involving representatives from national and county governments, public health institutions, technical partners, and emergency response practitioners. Feedback from the validation process informed revisions to decision pathways, escalation criteria, notification requirements, and response guidance prior to implementation. The validated DMT-PHE was designed to link with existing KNPHI surveillance, incident management, and PHEOC decision flows by standardizing how events are assessed, classified, notified, escalated, and activated for response. It subsequently served as the authoritative knowledge source for development of the DMT-PHE AI Agent, with all decision logic, escalation pathways, and response recommendations derived directly from the framework and its supporting technical guidance(21,26)

### Pilot Evaluation Design

This study represents the pilot evaluation phase of this broader development process. A mixed-methods evaluation was conducted, combining quantitative assessment of technical performance, usability, and acceptability with qualitative participant feedback. The evaluation employed standardized simulated outbreak scenarios to assess the ability of the DMT-PHE AI Agent to generate recommendations consistent with expert-defined DMT-PHE classifications and response pathways before routine operational deployment.

### AI System Architecture and Knowledge Base

The DMT-PHE AI Agent was developed as an artificial intelligence-enabled decision-support system to operationalize the validated Decision-Making Tool for Public Health Emergencies (DMT-PHE). The system was designed to transform a static decision framework into an interactive decision-support platform capable of assisting public health practitioners with event assessment, classification, escalation, notification, and identification of recommended response actions while maintaining human oversight and alignment with nationally approved public health emergency management procedures (9,22–25,30)

The AI Agent employs a retrieval-augmented generation (RAG) architecture that combines the reasoning capabilities of a large language model (Anthropic Claude Sonnet 4.5) with a curated, domain-specific knowledge base. Rather than relying solely on information embedded within the foundation model, the retrieval component identifies relevant content from authoritative reference documents before generating responses. This approach enables recommendations to be grounded in validated public health guidance, reduces dependence on parametric model knowledge, and improves the transparency and contextual relevance of generated outputs (22,30–32)

The knowledge base comprised a version-controlled collection of reference materials derived from the validated DMT-PHE framework together with supporting national and international guidance. These resources included the International Health Regulations (2005), Kenya public health emergency management policies, Public Health Emergency Operations Centre (PHEOC) procedures, disease-specific technical guidance, One Health frameworks, and the DMT-PHE Trigger Cards that provide standardized operational guidance for priority public health hazards (1,13,21,26–29) . For the purposes of evaluation, the knowledge base remained static throughout the study to ensure that all participants interacted with an identical corpus of reference materials and decision logic, thereby supporting reproducibility of the evaluation. Internet search functionality was disabled during testing to ensure consistency of outputs across participants.

When a user submitted an event description, the AI Agent interpreted the information, retrieved the most relevant guidance documents, and synthesized the retrieved evidence to generate structured recommendations. Outputs included event classification, escalation level, notification pathways, priority response actions, coordination mechanisms, and supporting references where available. Recommendations were generated using a hierarchical decision logic that prioritized the DMT-PHE framework, followed by nationally approved public health emergency management guidance, International Health Regulations (2005) requirements, and One Health guidance where applicable (1,13,21,27–29)

Where multiple knowledge sources addressed the same decision point, recommendations were resolved using a predefined hierarchy in which the validated DMT-PHE framework served as the primary authority. National public health emergency management policies were given precedence over international guidance where operational procedures differed, while the International Health Regulations (2005) and One Health guidance provided complementary decision support for notification, multisectoral coordination, and risk assessment. This approach ensured consistency with Kenya’s public health emergency management architecture while maintaining alignment with international best practices (1,22,24)

The DMT-PHE AI Agent was intentionally designed as a human-in-the-loop decision-support system rather than an autonomous decision-maker (Figure 1). Users retained responsibility for interpreting recommendations and making final operational decisions. The architecture also supports future updating of the knowledge base as national policies, disease-specific guidance, and international recommendations evolve, thereby facilitating continuous improvement while preserving transparency, traceability, governance, and human oversight. (9,10,22,23)

**Figure 1:**
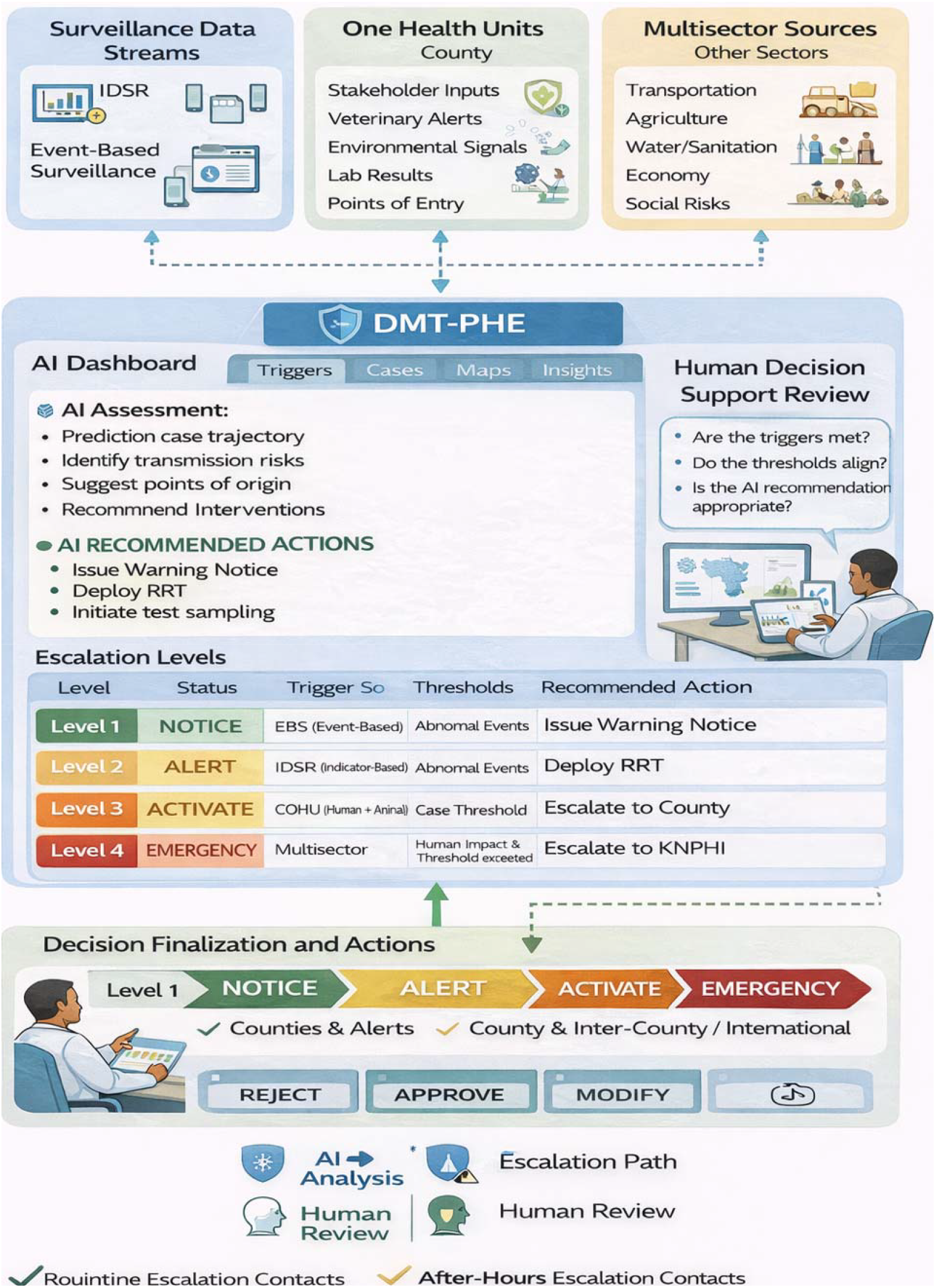
AI-enabled DMT-PHE Agentic architecture, multi-stream input interface, and escalation workflow

### Evaluation framework

The evaluation was designed to assess the technical performance, safety, usability, and acceptability of the DMT-PHE AI Agent during the pre-deployment phase using standardized simulation scenarios. The evaluation framework was informed primarily by the DECIDE-AI reporting guideline for early-stage assessment of AI-enabled decision-support systems, which emphasizes transparent evaluation of AI interventions prior to routine implementation (25). Additional reporting considerations were drawn from the CONSORT-AI and SPIRIT-AI extensions to promote comprehensive documentation of the AI intervention and its evaluation methodology (33,34).

Consistent with DECIDE-AI recommendations, the evaluation examined multiple domains of system performance, including agreement between AI-generated recommendations and expert-defined gold standards, completeness of recommended response actions, citation performance, safety, usability, and user acceptability. Because the evaluation was conducted using standardized simulated outbreak scenarios rather than live operational deployment, the study focused on feasibility and early-stage performance rather than effectiveness under routine public health practice (25,33,34). The evaluation further incorporated principles for trustworthy artificial intelligence, including transparency, explainability, human oversight, governance, and accountability (22–24)

Performance was assessed by comparing AI-generated recommendations against predefined gold-standard responses developed by subject matter experts. Operational relevance was evaluated through assessment of response completeness and contextual appropriateness. Usability and user experience were measured using standardized questionnaires, while safety was assessed through review of outputs for potentially harmful or inappropriate recommendations.

### Study participants recrutiment and orientation

Participants were purposively recruited from the Kenya National Public Health Institute (KNPHI) and selected county surveillance teams to ensure representation of professionals routinely involved in public health emergency preparedness and response. Eligible participants included epidemiologists, disease surveillance officers, public health emergency management personnel, and other technical staff with responsibilities related to event detection, risk assessment, outbreak investigation, or emergency coordination.

Eleven public health professionals participated in the pilot evaluation. Participants represented both national and subnational public health systems and had prior experience with infectious disease surveillance, outbreak response, implementation of the International Health Regulations (2005), and public health emergency management. Their expertise ensured that evaluation of the AI Agent reflected the perspectives of intended end-users responsible for operational decision-making.

Participation was voluntary, and all participants received a standardized orientation on the objectives of the evaluation and procedures for interacting with the DMT-PHE AI Agent before commencement of the simulation exercises. The orientation focused on familiarization with the evaluation process rather than training participants on interpretation of the outbreak scenarios or expected responses, thereby minimizing potential bias in the assessment.

### Development of Evaluation Scenarios

Three standardized outbreak scenarios were developed to evaluate the performance of the DMT-PHE AI Agent across distinct public health emergency contexts. Scenario selection was informed by the hazard categories represented within the DMT-PHE framework and designed to evaluate the system’s ability to apply decision logic under differing epidemiological and operational circumstances.

The scenarios comprised:

1. Public Health Event of Initially Unknown Etiology (PHEIUE), representing an unusual event requiring rapid assessment under conditions of uncertainty;
2. Rift Valley fever (RVF), representing a zoonotic outbreak requiring multisectoral coordination and application of One Health principles; and
3. Mpox, representing a directly transmitted infectious disease requiring assessment of escalation, notification, and response measures.

Scenario narratives were developed using realistic epidemiological information based on public health surveillance reports and outbreak investigation principles. Each scenario included sufficient information for participants to assess event characteristics while maintaining a standardized level of complexity across all evaluation sessions. Collectively, the three scenarios were selected to challenge different components of the DMT-PHE decision logic, including event classification, escalation pathways, notification requirements, multisectoral coordination, and recommended response actions (1,29). Each participant independently interacted with the DMT-PHE AI Agent using all three scenarios, resulting in a total of thirty-three scenario evaluations.

### Development of gold-standard responses

For each scenario, gold-standard responses were derived a priori by applying the DMT-PHE classification criteria directly to the scenario parameters which allowed unambiguous assignment of the expected escalation level. Gold-standard responses were derived from the validated DMT-PHE framework (21,26)

### Evaluation procedures

Participants received a standardized briefing on the objectives of the evaluation, study procedures, and use of the DMT-PHE AI Agent prior to commencement of the exercise. They then independently evaluated the AI Agent using each of the three standardized outbreak scenarios in a controlled simulation environment. Scenario information was presented in a uniform format to ensure consistency across participants. Participants were instructed to review each scenario, enter the information into the DMT-PHE AI Agent through its conversational interface, and assess the recommendations generated as they would during routine public health practice.

AI-generated outputs were documented using a structured electronic evaluation instrument developed in KoboToolbox(35). The instrument captured participants’ assessment of multiple performance domains, including accuracy of event classification, appropriateness of escalation recommendations, completeness of response actions, relevance of recommendations, citation support, trustworthiness of outputs, system usability, and potential safety concerns. Participants also completed a structured usability and acceptability questionnaire and provided qualitative feedback on the functionality, strengths, limitations, and potential operational applications of the system. All evaluations were completed independently to minimize influence from facilitators or other participants. Responses generated by the DMT-PHE AI Agent were subsequently compared with the predefined gold-standard responses using standardized evaluation criteria.

### Evaluation Outcome measures

The primary outcome was concordance between AI-generated recommendations and expert-defined gold-standard classifications for each outbreak scenario. Escalation concordance was scored on a three-point ordinal weighting scheme: 1.0 for exact agreement with the gold-standard level, 0.5 for an adjacent level, and 0 for a non-adjacent level or no level reported. The mean concordance score was the average across interactions; exact agreement was reported separately as the proportion scoring 1.0.

Concordance was assessed by determining whether the AI Agent correctly identified the expected event classification and corresponding escalation level defined within the validated DMT-PHE framework.

Secondary outcomes included:

- Response action coverage: Assessment of whether the AI Agent identified the priority response actions specified within the predefined gold standards, including notification requirements, coordination mechanisms, and key public health interventions.
- Citation performance: Assessment of the presence, appropriateness, and relevance of supporting references provided by the AI Agent. Citations were considered satisfactory when they appropriately supported the recommendations generated. Absence of citations was recorded separately from incorrect or misleading citations.
- Safety assessment: Identification of outputs that could potentially result in inappropriate public health actions, delayed escalation, incorrect notification pathways, omission of critical response actions, or recommendations inconsistent with the validated DMT-PHE framework. Safety observations were independently reviewed against the predefined gold standards and categorized according to their potential operational significance.
- Usability: System usability was assessed using the System Usability Scale (SUS), a validated ten-item instrument widely used for evaluation of digital health technologies. SUS scores range from 0 to 100, with higher scores indicating greater usability [25,26].
- User acceptability: Participants rated perceived usefulness, trustworthiness, contextual relevance, anticipated efficiency gains, and overall willingness to use the DMT-PHE AI Agent in routine public health practice using structured Likert-scale questions.

Qualitative feedback collected through open-ended questions was analyzed to identify common themes regarding usability, perceived strengths, limitations, and recommendations for future implementation.

### Statistical Analysis

Quantitative analyses were performed in R and were descriptive. Wilson 95% confidence intervals were reported for the principal proportions. The analyses reflected the exploratory objectives of this pilot evaluation. Participant characteristics, concordance scores, response action coverage, citation performance, safety observations, and usability measures were summarized using frequencies, percentages, means, and standard deviations where appropriate.

System Usability Scale scores were calculated according to the standardized scoring methodology described by Brooke, with interpretation based on established adjective rating scales proposed by Bangor and colleagues (36,37). Responses to user acceptability questions were summarized using descriptive statistics, including mean Likert-scale scores.

Qualitative responses were reviewed using a thematic descriptive approach. Participant comments were independently examined and grouped into recurring themes relating to perceived usefulness, strengths, limitations, trust, transparency, and potential operational applications of the DMT-PHE AI Agent. Given the exploratory nature of the study and limited sample size, no formal inferential hypothesis testing was undertaken.

### Ethical considerations

The evaluation was conducted using standardized simulated public health emergency scenarios and was designed to assess the DMT-PHE AI Agent under controlled conditions without influencing actual public health decisions. Participation by public health professionals was voluntary, and all evaluation data were anonymized prior to analysis and reported in aggregate form to protect participant confidentiality. The AI Agent functioned solely as a decision-support tool, with participants retaining responsibility for interpreting recommendations. The evaluation was conducted in accordance with principles of responsible AI, including transparency, explainability, human oversight, accountability, and governance (22–25)

## Results

### Participant characteristics

All eleven (11) enrolled public health professionals completed the evaluation, each independently assessing the three standardized outbreak scenarios resulting in 33 individual scenario evaluations (11 participants × 3 scenarios). Characteristics are shown in Table 1

**Table 1:** Participant characteristics (N = 11).

| Characteristic | Category | n (%) |
| --- | --- | --- |
| Role | Epidemiologist | 4 (36.4) |
|  | Disease surveillance officer | 4 (36.4) |
|  | Public health officer | 2 (18.2) |
|  | Emergency response officer | 1 (9.1) |
| Experience | 2–5 years | 6 (54.5) |
|  | 6–10 years | 2 (18.2) |
|  | >10 years | 3 (27.3) |
| Prior DMT-PHE use | Used in training | 8 (72.7) |
|  | Seen but not used | 2 (18.2) |
|  | Used in a real emergency | 1 (9.1) |
| Prior AI tool use | Yes, regularly | 9 (81.8) |
|  | Yes, a few times | 2 (18.2) |

### Overall System Performance

Escalation recommendations matched the gold-standard level exactly in 29 of 33 interactions (87.9%, 95% CI 72.7–95.2); the mean concordance score for event classification, giving partial credit for adjacent-level event recommendations, was 92.4%. The four non-exact responses were adjacent-level classifications, except one Scenario 3 interaction in which no level was returned (Figure 2). The three adjacent-level misses were under-escalations by one level (Level 2 against a Level 3 gold standard), with the same disease surveillance officer under-escalating in both Scenario 1 and Scenario 2; the Scenario 3 miss was a failure to return a level rather than a misclassification.

**Figure 2:**
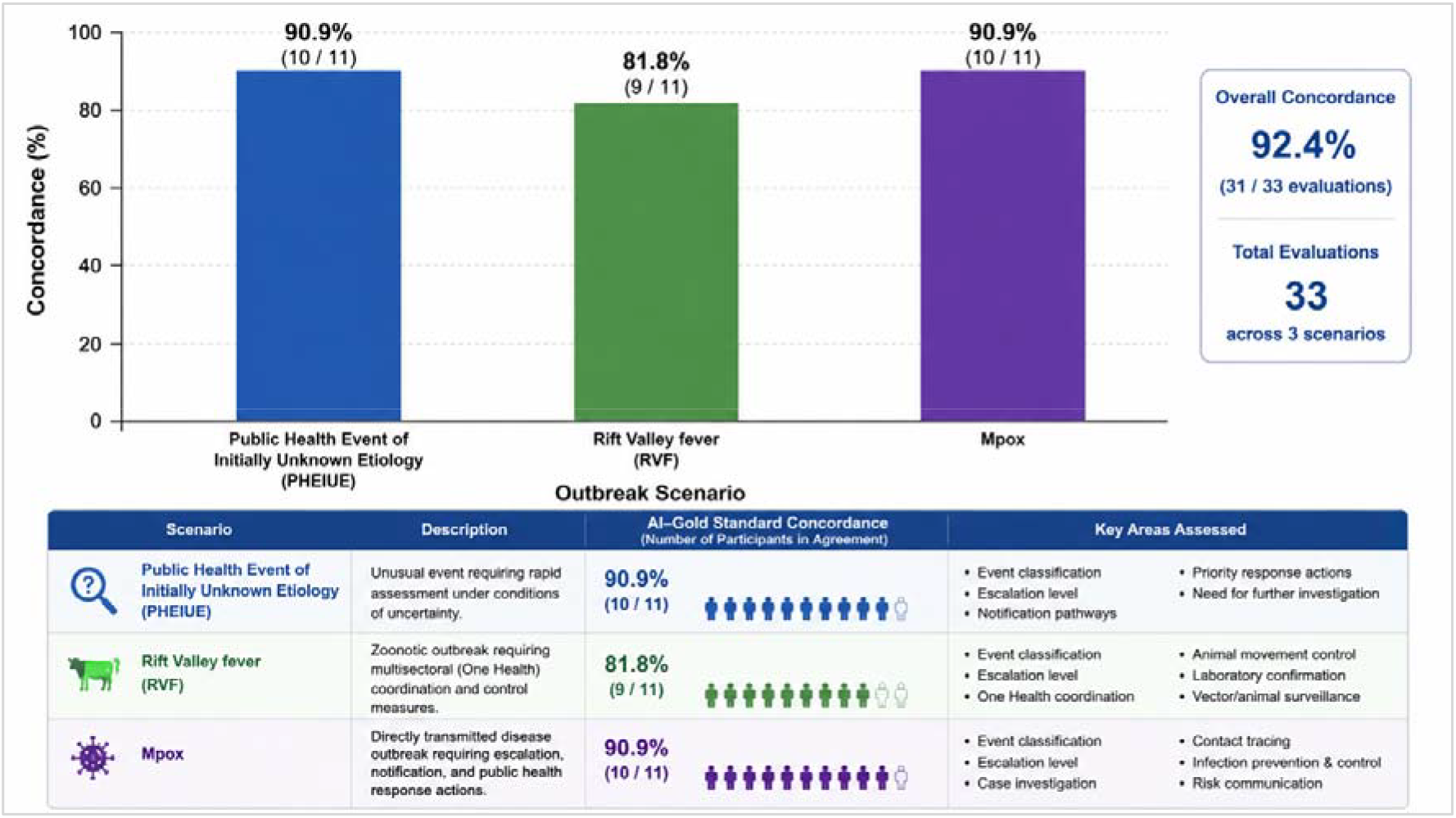
Scenario-specific concordance of AI-generated reccomendations Note: Concordance is the proportion of AI generated event classifications and escalation reccommendations that agreed with expert defined gold-standard reference

### Scenario-specific concordance

Performance was consistently high across the three outbreak scenarios, although modest variation was observed according to event type. For the Public Health Event of Initially Unknown Etiology (PHEIUE) scenario, the AI Agent achieved 90.9% concordance with the predefined gold-standard responses. For the Rift Valley fever (RVF) scenario, concordance was 81.8%. The AI Agent appropriately identified the zoonotic nature of the event and consistently recommended multisectoral coordination, laboratory confirmation, and implementation of One Health response measures. For the Mpox scenario, concordance returned to 90.9%, with the AI Agent correctly identifying the event classification, escalation requirements, notification pathways, and priority response measures in the majority of evaluations. Scenario-specific concordance is summarized in Figure 2

### Coverage of recommended response actions

Coverage was high for core notification and One Health actions but variable for others. WHO/IHR notification was identified in 45.5% of PHEIUE interactions, PPE/IPC in 45.5% of Rift Valley fever interactions, the 21-day Mpox contact-tracing period in 9.1%, and specific exposed-health-worker guidance in 27.3% (Table 2).

**Table 2:** Coverage of pre-specified key response actions by scenario (N = 11).

| Scenario | Pre-specified action | Mentioned, n/11 (%) |
| --- | --- | --- |
| S1 — PHEIUE, Busia | KNPHI notification | 11 (100.0) |
|  | PPE and IPC measures | 10 (90.9) |
|  | Cross-border coordination with Uganda | 9 (81.8) |
|  | WHO notification / IHR obligations | 5 (45.5) |
| S2 — RVF, Garissa/Tana River | One Health / veterinary engagement | 11 (100.0) |
|  | Animal movement restriction | 11 (100.0) |
|  | KNPHI notification | 10 (90.9) |
|  | PPE and IPC measures | 5 (45.5) |
| S3 — Mpox, Embu | KNPHI notification | 11 (100.0) |
|  | Healthcare-worker exposure addressed (any) | 10 (90.9) |
|  | Healthcare-worker exposure, specific guidance | 3 (27.3) |
|  | Contact tracing period specified (21 days) | 1 (9.1) |

### Citation performance

The assistant provided at least one DMT-PHE citation in 26 of 33 interactions (78.8%). Of 51 adjudicated citations, 24 (47.1%) were Correct and 27 (52.9%) Partially correct (relevant DMT-PHE content but not the primary authoritative reference); none was Incorrect or Unverifiable, and the assistant drew on a narrow, reproducible set of pages (Table 3). In the seven evaluations that generated recommendations without explicit citation of supporting references, the responses remained consistent with the predefined gold-standard recommendations and did not result in incorrect classification, escalation, or response guidance.

**Table 3:** Citation presence and accuracy by scenario.

| Scenario | Interactions with ≥1 citation, n/11 (%) | Citations adjudicated, n | Correct, n (%) | Partially correct, n (%) | Incorrect/Unverifiable, n |
| --- | --- | --- | --- | --- | --- |
| S1 — PHEIUE, Busia | 9 (81.8) | 18 | 9 (50.0) | 9 (50.0) | 0 |
| S2 — RVF, Garissa/Tana River | 8 (72.7) | 16 | 8 (50.0) | 8 (50.0) | 0 |
| S3 — Mpox, Embu | 9 (81.8) | 17 | 7 (41.2) | 10 (58.8) | 0 |
| Overall | 26 (78.8) | 51 | 24 (47.1) | 27 (52.9) | 0 |

### Safety Assessment

Safety observations were recorded in 4 of 33 interactions (12.1%); two were formally adjudicated and both coded Uncertain, No AI-generated recommendation was classified as representing a significant safety concern. Specifically, no outputs were identified that would have resulted in inappropriate event classification, incorrect escalation, omission of critical notification pathways, or recommendations likely to delay or compromise outbreak response. Verbatim agent outputs were not captured, which limited adjudication (see Limitations). Detailed observations are provided in Supplementary.

### System Usability and User Acceptability

The mean SUS score was 85.2 (SD 11.4, range 67.5–100), indicating excellent usability with 7 of 11 participants scoring Excellent and none Poor. User acceptability ratings were positive, Participants reported high levels of trust in the recommendations generated by the AI Agent (mean 4.27/5), perceived strong contextual relevance to public health emergency management (4.55/5), and anticipated substantial time savings compared with manual consultation of guidance documents (4.82/5). Overall willingness to use the system during routine public health emergency management activities was also high (Table 4).

**Table 4:** Usability and acceptability (N = 11).

| Domain | Measure | Result |
| --- | --- | --- |
| System Usability (SUS) | Mean (SD) | 85.2 (11.4) |
|  | Range | 67.5–100 |
|  | Grade: Excellent / Good / Adequate / Poor | 7 / 2 / 2 / 0 |
| Acceptability (mean, SD) | Trust to act on recommendations | 4.27 (0.65) |
|  | Relevance to Kenyan PHE context | 4.55 (0.52) |
|  | Time saving vs manual lookup | 4.82 (0.40) |
|  | Concern about harmful guidance<sup>†</sup> | 2.09 (0.83) |
|  | Would recommend to colleagues | 4.36 (1.29) |

<sup>†</sup> Higher score indicates greater concern.
SUS scored 0–100; acceptability items scored 1–5.

### Qualitative feedback

Open-text responses (8 to 10 of 11 participants) most consistently praised the agent’s speed and conciseness; one epidemiologist described the outputs as “sharp and brief and not generalist.” Improvement suggestions fell into three areas: broader source coverage beyond the DMT-PHE (two participants), greater contextual depth (one epidemiologist found responses “highly summarised”), and offline or mobile access for staff in hard-to-reach areas (one emergency response officer). Participants saw the clearest use case where subject-matter experts are not immediately reachable and real-time guidance is needed. They considered the AI Agent particularly valuable for supporting less experienced public health practitioners and strengthening standardized implementation of the DMT-PHE framework across counties.

## Discussion

This study describes the development and pilot evaluation of the Decision-Making Tool for Public Health Emergencies (DMT-PHE) AI Agent, an artificial intelligence-enabled decision-support system designed to operationalize a nationally validated public health emergency decision framework. The AI Agent was developed through an iterative, multidisciplinary process that translated established decision pathways, escalation criteria, and response protocols into an interactive, explainable platform capable of supporting operational public health decision-making. By embedding validated national guidance within a structured AI architecture, the system was designed to augment, rather than replace, professional judgement while improving the accessibility and consistent application of public health emergency guidance during rapidly evolving events.

This pilot evaluation demonstrated that the DMT-PHE AI Agent achieved high concordance with expert-defined public health emergency classifications and escalation decisions while demonstrating excellent usability and strong user acceptance. Across 33 standardized scenario evaluations, the AI Agent correctly applied the decision logic embedded within the validated DMT-PHE framework, consistently identified key response actions, and generated recommendations without major safety concerns. Participants also reported high levels of trust, contextual relevance, and anticipated efficiency gains, suggesting that the system was both technically robust and operationally acceptable to its intended users.

These findings are encouraging because they address an important but underexplored gap in public health emergency management. Considerable investments have strengthened surveillance, laboratory systems, and epidemic intelligence over the past two decades; however, less attention has been given to supporting the operational decisions that occur after potential threats have been detected. Translating surveillance information into timely and appropriate public health action remains dependent on the ability of practitioners to interpret multiple guidance documents, assess risk, determine escalation pathways, and coordinate response activities under conditions of uncertainty (1,4–6) The high concordance observed in this study suggests that AI-assisted decision support can help standardize these processes while remaining aligned with nationally validated guidance.

Most published applications of artificial intelligence in public health have focused on disease surveillance, outbreak prediction, epidemic intelligence, and forecasting (6–8). Comparatively little evidence exists on the use of AI to support operational public health emergency management functions such as event classification, escalation, notification, and response coordination. The DMT-PHE AI Agent differs from these approaches by operationalizing an existing national decision framework rather than generating predictions or replacing professional judgement.

The use of a retrieval-augmented architecture linked to a validated knowledge base also distinguishes the system from general-purpose conversational AI. Recommendations were grounded in nationally approved guidance, thereby improving transparency, traceability, and consistency while reducing reliance on model-generated responses alone (22,23,30,31). This approach aligns with recent WHO guidance advocating for AI systems that augment human expertise, operate within established governance structures, and provide explainable outputs for health decision-making(22,23).The DMT-PHE AI Agent therefore represents an example of how generative AI can be integrated into existing public health systems without displacing professional accountability.

Although this evaluation was conducted under simulated conditions, the findings suggest several opportunities for future implementation. Integration of the DMT-PHE AI Agent within KNPHI workflows, county surveillance systems, Public Health Emergency Operations Centres (PHEOCs), and event-based surveillance platforms could provide structured support for event assessment and escalation while promoting more consistent application of national guidance. Such support may be particularly valuable in decentralized settings where access to experienced public health emergency specialists is limited.

Participants also perceived that the AI Agent reduced the need to consult multiple guidance documents and could facilitate more rapid decision-making during outbreak investigations. While this study did not evaluate outbreak timeliness or operational outcomes, these observations suggest a plausible mechanism through which AI-assisted decision support could contribute to more timely implementation of public health actions and future achievement of outbreak performance targets such as the 7-1-7 framework (4,5,38). Demonstrating such impact, however, will require prospective evaluations conducted under routine operational conditions.

The findings further highlight the importance of maintaining human oversight during AI-supported decision-making. Public health emergency management requires consideration of epidemiological, operational, legal, political, and contextual factors that extend beyond algorithmic reasoning. Consequently, AI should be viewed as a decision-support tool that complements, rather than replaces, professional judgement. Appropriate governance, workforce training, and continuous updating of the knowledge base will remain essential for safe and sustainable implementation (22–24).

Although the findings demonstrate that the DMT-PHE AI Agent can accurately operationalize a validated public health emergency decision framework under simulated conditions, the study was not designed to determine whether AI-assisted decision support improves outbreak detection, response activation, or achievement of operational performance targets such as the 7-1-7 framework (4,5). Demonstrating these outcomes will require prospective implementation studies conducted under routine public health practice. Furthermore, successful deployment will depend not only on technical performance but also on appropriate governance, workforce development, and sustained human oversight. Consistent with international guidance on trustworthy AI, mechanisms for routine validation, knowledge-base updating, accountability, and user training will be essential to minimize automation bias and ensure that the DMT-PHE AI Agent remains a decision-support tool that complements, rather than replaces, professional judgement (22,23,25,30–32,39,40)

### Strengths, Limitations and Future Directions

This study has several important strengths. To our knowledge, it represents one of the first structured evaluations of an artificial intelligence-enabled decision-support system specifically developed to operationalize a nationally validated public health emergency decision framework. Unlike many AI applications that focus on disease prediction or surveillance, the DMT-PHE AI Agent was designed to support operational decision-making by translating established guidance into structured recommendations for event classification, escalation, notification, and response. The evaluation employed expert-developed gold standards derived from the validated DMT-PHE framework, incorporated multiple outbreak scenarios representing diverse public health contexts, and assessed not only technical performance but also safety, usability, citation performance, and user acceptability using an internationally recognized evaluation framework (21,25,26,33,34)

Several limitations should also be acknowledged. First, the evaluation included a relatively small group of experienced public health professionals who routinely participate in outbreak preparedness and response, and findings may not be generalizable to less experienced users or to other health systems. Second, the use of standardized simulation scenarios cannot fully replicate the uncertainty, incomplete information, competing priorities, and operational pressures encountered during real-world public health emergencies. Third, participants interacted with the DMT-PHE AI Agent during a single evaluation session, precluding assessment of changes in user behaviour, trust, or learning over time. Although AI-generated outputs were assessed using structured evaluation instruments, verbatim system responses were not systematically archived for independent review, limiting opportunities for detailed qualitative analysis of model reasoning.

The evaluation was also limited to three outbreak scenarios and therefore does not establish performance across the full range of hazards addressed by the DMT-PHE framework, including chemical, radiological, environmental, and complex humanitarian emergencies. Furthermore, because the AI Agent derives its recommendations from a curated knowledge base, its performance depends on the completeness, quality, and currency of the underlying guidance. Maintaining an up-to-date knowledge base will therefore be essential as national policies, disease-specific guidance, and international recommendations evolve. Finally, although no incorrect citations or unsafe recommendations were identified during this pilot evaluation, large language models remain susceptible to factual inaccuracies and automation bias. Continued monitoring, periodic validation, and sustained human oversight will remain essential to ensure that AI-supported recommendations remain accurate, transparent, and operationally appropriate (22,25,30,32,39,41)

Future research should evaluate the DMT-PHE AI Agent under routine operational conditions to determine whether AI-assisted decision support improves the consistency, timeliness, and quality of public health emergency management. Prospective implementation studies should assess integration with national surveillance systems, event-based surveillance platforms, PHEOCS, and county-level emergency management workflows while examining impacts on workflow efficiency, decision consistency, user behaviour, and outbreak performance indicators, including the 7-1-7 framework. Additional research should expand evaluation to a broader range of public health hazards, assess long-term maintenance of the knowledge base, explore adaptation of the framework to other country contexts, and examine the cost-effectiveness and implementation requirements of AI-assisted decision support within national public health systems (4,5,27,38,42)

## Conclusion

This study demonstrates the feasibility of translating a nationally validated public health emergency decision framework into an artificial intelligence-enabled decision-support system for operational public health practice. By operationalizing the Decision-Making Tool for Public Health Emergencies (DMT-PHE), the DMT-PHE AI Agent transforms complex national guidance into structured, transparent, and context-specific recommendations that support event assessment, classification, escalation, notification, and response while maintaining human oversight and alignment with established public health policies.

Beyond evaluating an AI application, this study provides evidence that artificial intelligence can be responsibly integrated into public health emergency management to augment professional judgement rather than replace it. The DMT-PHE AI Agent illustrates how validated national guidance can be converted into operational decision intelligence that is accessible, explainable, and aligned with routine public health workflows.

As countries continue to strengthen epidemic intelligence and emergency preparedness capacities, this work provides a proof of concept that nationally validated public health emergency decision frameworks can be successfully operationalized through responsible artificial intelligence. Such approaches offer a practical pathway for enhancing the consistency, transparency, and quality of public health emergency decision-making while preserving professional accountability and national ownership of decision processes.

## Data Availability

The datasets generated and analyzed during the current study are available from the corresponding authors upon reasonable request. Evaluation materials, including outbreak scenarios, facilitator guides, gold-standard responses, and assessment instruments, may be made available subject to approval by the Kenya National Public Health Institute and collaborating institutions.

## Declarations

### Consent to participate and Study Approval

This study was conducted using standardized simulated public health emergency scenarios and did not constitute human subjects research because it did not involve patients, biological specimens, invasive procedures, or identifiable personal data. Participation by public health professionals was voluntary, and informed consent was obtained prior to participation.

The study was conducted under administrative approval and clearance from the Kenya National Public Health Institute as it was determined to constitute a public health program evaluation and pre-deployment assessment and quality improvement of the DMT-PHE AI Agent to strengthen public health emergency preparedness and response.

### Authors’ Contributions

**Mark Nanyingi (MN):** Conceptualization of DMT-PHE, methodology, DMT-PHE framework development, evaluation design, interpretation of findings and manuscript drafting

**Eric Osoro (EO):** Conceptualization of evaluation methodology and design,,data curation, formal analysis, interpretation of findings, and manuscript review.

**Geoffrey H. Siwo (GS):** Conceptualization of DMT-PHE AI agent, artificial intelligence system development, methodology, technical oversight, interpretation of findings, and manuscript review.

**Isaac Ngere (IN):** Artificial intelligence system development support, evaluation design, technical validation, data interpretation, and manuscript review.

**Samuel Kadivane (SK):** Evaluation implementation, participants coordination, data collection, interpretation of findings, and manuscript review.

**James Magige (JM):** Evaluation implementation, data collection, interpretation of findings, and manuscript review.

**Shreya Jain (SJ):** Artificial intelligence system development, web platform, interpretation of findings, and manuscript review.

**Joseph Kamau (JK):** Validation and evaluation support, One Health technical input, interpretation of findings, and manuscript review.

**Bryan Nyawanda (BN):** Evaluation methodology, formal analysis, interpretation of findings, and manuscript review.

**Joseph Njoroge (JN):** Administrative support, and manuscript review.

Ian Njeru (IAN): Public health emergency management expertise, interpretation of findings, and manuscript review.

**Kadondi Kasera (KK):** Conceptualization of DMT-PHE, Evaluation oversight, interpretation of findings, and manuscript review.

**Victoria Kanana (VK):** Conceptualization of DMT-PHE, Validation and Evaluation oversight, interpretation of findings, and manuscript review.

**Maureen Kamene (MK):** Conceptualization of DMT-PHE, Supervision, public health emergency management oversight, interpretation of findings, and manuscript review. All authors reviewed, revised, and approved the final manuscript.

## Acknowledgements

The authors acknowledge the Kenya National Public Health Institute (KNPHI), participating county surveillance teams, and public health professionals who contributed to the pilot evaluation of the DMT-PHE AI Assistant. We further acknowledge the collaborative efforts of national and international partners involved in development of the DMT-PHE, whose work provided the foundation for the AI-assisted decision support system evaluated in this study.

